# Brainstem evoked auditory potentials in tinnitus: a best-evidence synthesis and meta-analysis

**DOI:** 10.1101/2022.01.29.22270068

**Authors:** Laura Jacxsens, Joke De Pauw, Emilie Cardon, Annemarie van der Wal, Laure Jacquemin, Annick Gilles, Sarah Michiels, Vincent Van Rompaey, Marc J.W. Lammers, Willem De Hertogh

**Affiliations:** Department of Rehabilitation Sciences and Physiotherapy, Faculty of Medicine and Health Sciences, University of Antwerp, Antwerp, Belgium; Department of Otorhinolaryngology, Head and Neck Surgery, Antwerp University Hospital (UZA), Edegem, Belgium; Department of Translational Neuroscience, Faculty of Medicine and Health Sciences, University of Antwerp (UA), Antwerp, Belgium; Department of Orofacial Pain and Dysfunction, Academic Centre for Dentistry Amsterdam (ACTA), University of Amsterdam, Amsterdam, the Netherlands; Department of Education, Health and Social Work, University College Ghent, Ghent, Belgium; Faculty of Rehabilitation Sciences, REVAL, University of Hasselt, Hasselt, Belgium

**Keywords:** Tinnitus, Auditory Evoked Potentials, Systematic Review

## Abstract

**Introduction:** Accumulating evidence suggests a role of the brainstem in tinnitus generation and modulation. Several studies in chronic tinnitus patients have reported latency and amplitude changes of the different peaks of the auditory brainstem response, possibly reflecting neural changes or altered activity. The aim of the systematic review was to assess if alterations within the brainstem of chronic tinnitus patients are reflected in short- and middle-latency auditory evoked potentials (AEPs).

**Methods:** A systematic review was performed and reported according to the PRISMA guidelines. Studies evaluating short- and middle-latency AEPs in tinnitus patients and controls were included. Two independent reviewers conducted the study selection, data extraction, and risk of bias assessment. Meta-analysis was performed using a multivariate meta-analytic model.

**Results:** Twenty-seven cross-sectional studies were included. Multivariate meta-analysis revealed that in tinnitus patients with normal hearing, significantly longer latencies of auditory brainstem response (ABR) waves I (SMD = 0.66 ms, p < 0.001), III (SMD = 0.43 ms p < 0.001), and V (SMD = 0.47 ms, p < 0.01) are present. The results regarding possible changes in middle-latency responses (MLRs) and frequency-following responses (FFRs) were inconclusive.

**Discussion:** The discovered changes in short-latency AEPs reflect alterations at brainstem level in tinnitus patients. More specifically, the prolonged ABR latencies could possibly be explained by high frequency sensorineural hearing loss, or other modulating factors such as cochlear synaptopathy or somatosensory tinnitus generators. The question whether middle-latency AEP changes, representing subcortical level of the auditory pathway, are present in tinnitus still remains unanswered. Future studies should identify and correctly deal with confounding factors, such as age, gender and the presence of somatosensory tinnitus components.

## Introduction

Tinnitus, or ‘ringing in the ears’, is the conscious perception of an auditory sensation in the absence of a corresponding auditory source. It is a very common symptom with a prevalence of 10 to 15% in an adult population (1). This symptom is often associated with reduced quality of life and psychosocial wellbeing (2). There are many factors associated with the onset of tinnitus, the most common one being hearing loss (3, 4). Other possible triggering factors include ototoxic medications, head and neck trauma, temporomandibular dysfunctions, neck pain, neurological and psychological conditions (1).

Literature strongly suggests that the brainstem has a role in tinnitus generation and modulation, as well as in nonauditory comorbid conditions associated with tinnitus, such as neck disorders, anxiety, sleep disorders, difficulty concentrating, and depression (5). Animal studies have consistently shown disturbances in the level and patterns of spontaneous neural activity of brainstem auditory nuclei, linked with the onset of tinnitus. More specifically, these changes include increased spontaneous firing rates and bursting activity, which are both forms of hyperactivity, and increased neural synchrony (5-7). These disturbances are first found in the cochlear nucleus and inferior colliculus (8-11) and may be relayed to higher levels of the pathway (5).

On functional magnetic resonance imaging (fMRI) scans, increased resting state activity is also found in the auditory nuclei in the brainstem (12, 13). Multiple structures in the brainstem, including the cochlear nuclei and inferior colliculi, display abnormal function linked to tinnitus (12, 14, 15). It is important to remember that these brainstem structures send signals via multiple pathways to other brainstem and cortical regions, resulting in a cascade of changes directly associated with tinnitus generation (5).

Among clinical procedures to assess various levels of the auditory system, the most widely used involve auditory evoked potentials (AEPs) (16, 17). It is a technique that is used for the evaluation of neural activity in the auditory pathway, from cochlea to auditory cortex (18). AEPs are generally categorized in three classes according to their latency: short-, middle- and long-latency AEPs (S1 figure). Short-latency AEPs, often referred to as auditory brainstem responses (ABRs) (19), are generated at the first part of the auditory pathway, from the most distal portion of the auditory nerve to the brainstem. ABR peaks are labelled with roman numerals I-VI, of which wave I, III and V are most reliably recorded (20). Middle-latency AEPs, also referred to as middle-latency responses or MLRs, are believed to be generated in the thalamus, in subcortical regions and in the primary auditory cortex (21). MLRs consist of three positive (P0, Pa, Pb) and two negative peaks (Na, Nb) (19, 21). Long-latency AEPs are generally a product of the neocortex reflecting higher-order, cortical processing (22).

Additionally, the frequency-following response (FFR) is distinguished from other evoked potentials by precisely reflecting the neural processing of a sound’s acoustic features (23, 24). One way to interpret FFR responses is by examining the timing of response peaks in the time-domain waveform. By applying a fast Fourier transform (FFT), the encoding strength of individual frequencies in the FFR can be examined, such as the fundamental frequency (F_0_), the first formant (F_1_), and high harmonics (HH) (23). The FFR has a stimulus-to-response latency of 5-9 ms (25) and could therefore be considered as a short-latency AEP. This response is believed to be generated predominantly in the auditory midbrain (26-30), a hub of afferent and efferent activity (31). Consequently, the FFR reflects an array of influences from the auditory periphery and the central nervous system (23). FFR recordings are increasingly considered a valuable tool to index the current functional state of the auditory system (32).

The recently published systematic review and meta-analysis by Cardon et al. (33) provides an overview of the literature regarding long-latency AEPs in subjective tinnitus patients. A decreased amplitude and prolonged latency of P300 was observed, resulting in the consideration of this potential as a prospective biomarker for subjective tinnitus. This potential is mainly observed in the central and parietal regions of the cerebral cortex (34) and is often used as a measure of cognitive processing (35, 36).

There is no consensus yet on potential AEP changes at the level of the brainstem and the midbrain. Evidence from animal studies with salicylate-induced tinnitus revealed shorter ABR peak latencies, reduced wave I amplitudes, and increased amplitude of wave IV (37). In contrast, in animals with noise-induced tinnitus, all ABR waves had reduced amplitudes (37). This implicates that salicylate and noise induce different changes within the auditory brainstem, but still cause the tinnitus percept.

Since there is evidence suggesting a role of the brainstem in tinnitus generation, our aim was to perform a systematic review to examine if alterations in the brainstem auditory nuclei in tinnitus patients are reflected in short- and middle-latency AEPs. Based on experimental laboratory studies, we expect to find shorter peak latencies and larger amplitudes of the brainstem responses, reflecting increased neural synchrony.

## Materials and methods

### Protocol registration

The protocol of this study has been registered in PROSPERO on 04/05/2021 (ID 243687) at https://www.crd.york.ac.uk/PROSPERO/. The Preferred Reporting Items for Systematic Reviews and Meta-analyses Protocols (PRISMA-P) statement (38, 39) was the guideline during the design and writing of this study.

### Eligibility criteria

Regarding study population, adults with chronic subjective tinnitus were included. The following exclusion criteria were implemented: no tinnitus, objective tinnitus, pulsatile tinnitus, tinnitus caused by middle ear pathology, tinnitus caused by a tumor, brain tumors, sudden sensorineural hearing loss, drug induced tinnitus, Ménière’s disease, Schwannoma, healthy subjects, alcoholism, intracranial hypertension, multiple sclerosis, diabetes, cerebrovascular disease, Alzheimer’s disease, Parkinson’s disease, migraine. The included outcomes were all short- and middle-latency AEP measures; long-latency AEPs were excluded. As for study design, reviews, systematic reviews, and meta-analyses were excluded.

### Search strategy

The search strategy was based on the domain-determinant-outcome model. In this model, the domain was defined as adults with chronic subjective tinnitus. Short- and middle-latency AEPs were the determinants. Finally, the outcome was described as the prevalence of alterations in short-and middle-latency auditory evoked potentials in tinnitus patients compared to controls. The databases that were searched in the scope of this systematic review and meta-analysis are PubMed and Web of Science. Search strings were adapted for each of these databases. The search strategy included terms relating to tinnitus and short- and middle-latency auditory evoked potentials and has been evaluated by an independent librarian from the University of Antwerp, as is recommended by the Institute of Medicine (40). Only primary research published in English and Dutch was considered for this review. There were no restrictions on date of publication. Database searching ended on 30/04/2021. The search strategies for PubMed and Web of Science are presented in the appendix (S2).

### Study selection

Titles and abstracts of the articles retrieved by the database searches were screened by two independent authors (LJ and JDP). Articles that were included based on the title and abstract and met the eligibility criteria, were subsequently subjected to a full-text screening by the same two independent authors. In case of disagreement, this was resolved by a consensus meeting between the two reviewers. If a consensus could not be reached, an extra reviewer (WDH) was consulted.

### Data extraction

A standardized form was used for data extraction. The following data were extracted by the two reviewers (LJ and JDP): study design, study population (sample size, sex, age, hearing level), study protocol/methodology, outcome measures (methods of AEP measurements, AEP component(s), characteristics (latency, amplitude)), and results. If reported, measures on tinnitus duration, loudness, and subjective severity were also included in the data extraction tables.

### Risk-of-bias and quality assessment

Two reviewers (LJ and JDP) evaluated the quality of the studies independently based on a checklist. Disagreements between authors were solved by discussion or with a third reviewer (WDH). To assess the methodological quality of cross-sectional studies, the Joanna Briggs Checklist for Analytical Cross-Sectional Studies, which consists of eight items, was used. Each item was assessed as “yes,” “no,” “unclear” or “not applicable.” By analogy with Marshall et al. (41), we assigned a score of 1 to a “yes” rating for each of the 8 criteria, resulting in a score from 0-8. A cut-off score of 4 was used to exclude low-quality studies from synthesis. Moderate risk of bias was defined as a score of 5 or 6 and low risk of bias to scores of 7 and 8.

### Meta-analyses

Meta-analyses were conducted using the Metafor package in R (version 3.6.2, © 2019 The R Foundation for Statistical Computing) (42). Effect sizes were calculated as standardized mean differences between tinnitus groups and control groups. In order to minimize clinical variety and considering our main goal was to investigate the possible influence of tinnitus on AEPs, without hearing loss as a (possible) influencing factor, we only included papers which specified that the included tinnitus patients had clinically normal PTA thresholds (≤ 20 dB HL) in our meta-analyses. Papers in which tinnitus patients had other comorbidities, such as temporomandibular dysfunctions, were also excluded in the final meta-analysis. Data pooling was considered if studies were clinically homogeneous.

Since several included papers reported data on multiple short- and middle-latency AEP components within the same group of subjects, sampling errors of these results were expected to be correlated. To account for this correlation, a multivariate model was applied. Furthermore, AEP components needed to be reported in a minimum of three papers to be included in the meta-analysis. This is in analogy to Cardon et al. (33). In a multivariate meta-analysis, covariances between the sampling errors of various outcome measures are a necessary addition to the model. However, the correlations between several outcome measures within one paper, which is required information to compute these covariances, are often not reported. To account for this lack of information, a variance-covariance matrix was constructed based on correlations between different AEP components in a dataset used in our previously published study in which ABRs in young adults with and without tinnitus were acquired (33, 43).

In order to assess statistical heterogeneity in this multivariate model, forest plots were inspected and I^2^ was computed according to the approach described by Jackson et al. (44). This approach is based on the variance-covariance matrix of the fixed effects under the model with random effects and the model without. In order to explore outliers or influential studies, post hoc analyses were performed for all ABR components included in the multivariate model. Outlier detection was based on Cook’s distance and influence diagnostics were used to visualize influence of individual studies. The identified influential studies were not removed from the final analysis, since outliers and influential cases might reveal important patterns regarding study characteristics that could be acting as potential moderators (45). Furthermore, evidence for publication bias was investigated in using funnel plots and Egger’s regression tests.

## Results

### Study selection

In total, 1,209 articles were retrieved from the searched databases. After the removal of 313 duplicates, the articles went through a first screening phase based on title and abstract. This resulted in the exclusion of 829 articles. After full-text screening and critical appraisal, 27 papers were included. A detailed overview of the study selection process can be found in the PRISMA flowchart in figure 1.

**Figure 1.**
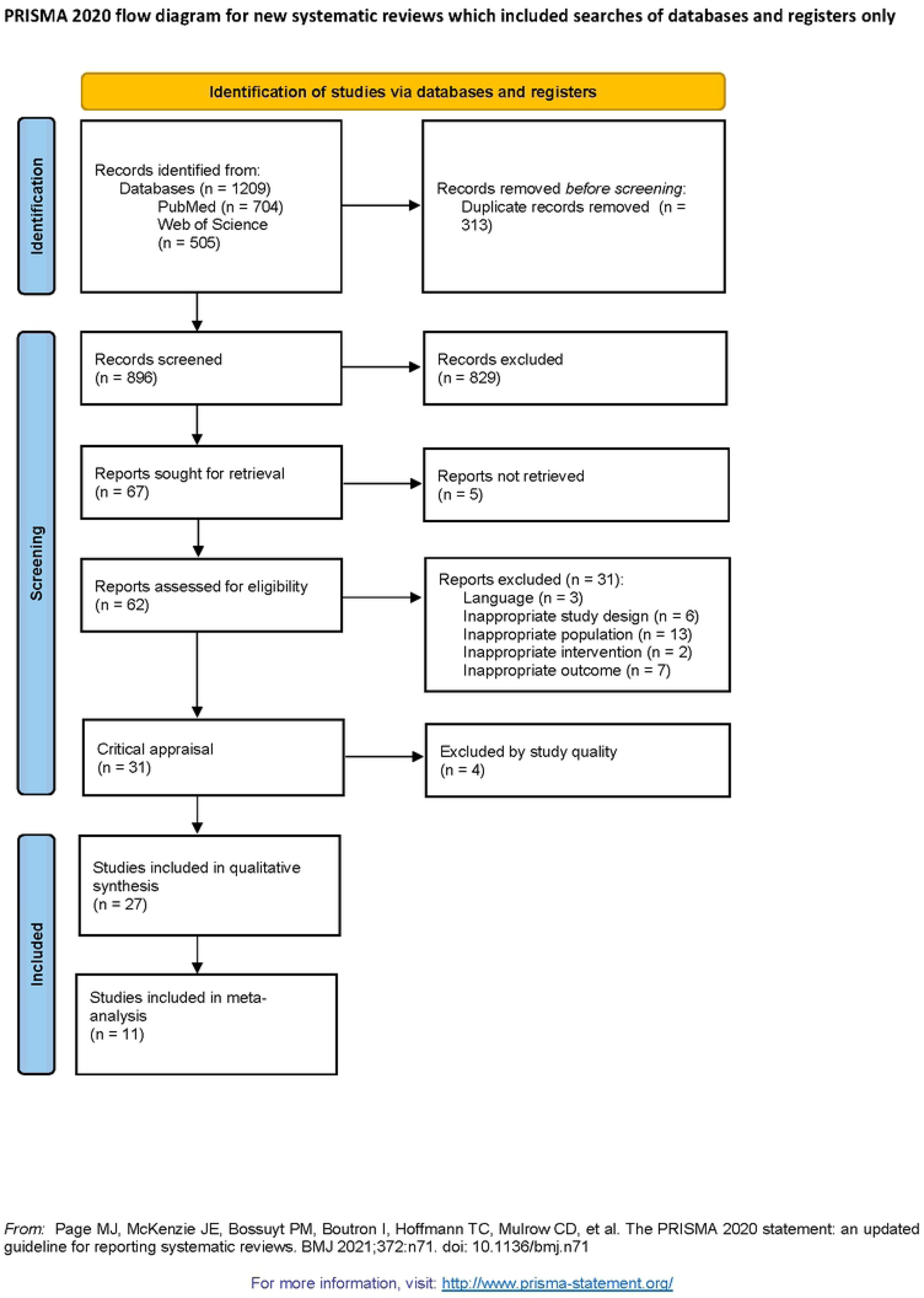
PRISMA flowchart of the study selection procedure.

### Study characteristics

Twenty-seven cross-sectional studies comparing AEPs between tinnitus patients and controls were included. The average number of tinnitus patients enrolled in these studies was 27, ranging from 10 to 113. On average, 35 control subjects, ranging from 10 to 220, were included. The mean age of tinnitus patients was 37.8 years, ranging from 18 to 68 years, and the mean age for controls was 34.2 years, ranging from 18 to 68 years (n = 24 papers). The proportion of male patients (reported in 23 studies) in the tinnitus group was, on average, 60.1% (ranging from 0% to 100%). In control subjects, the proportion of male subjects was 57.0% (ranging from 0% to 100%). The mean duration of tinnitus (reported in 8 studies) was 34 months.

The researched AEP varied across papers. In 24 studies, ABRs were measured, all of which used click stimuli to elicit the responses. The study by Pinkl et al. (46) used both click stimuli and tone burst stimuli. The most commonly studied ABR parameters were latencies of wave I (n = 21), wave III (n = 20), and wave V (n = 21). Interpeak latencies (IPLs) I-III (n = 14), III-V (n = 14), and I-V (n = 16), and amplitudes of waves I (n = 15), III (n = 11), and V (n = 16) were also frequently studied. Amplitude ratios III/I, V/III, and V/I; were only reported in 5, 2, and 8 papers respectively. MLRs (16, 47, 48) and FFRs (49-51) were acquired in three studies each.

For each individual study, a summary of the characteristics of the tinnitus group and control group, and main results are presented in the appendix (S3 tables). Different AEP components, more specifically ABRs, MLRs, and FFRs, were investigated in the different cross-sectional papers. The following sections go into more detail about each of these components.

### Risk of bias

The studies that met the inclusion criteria were assessed for risk of bias. According to our cutoff scores, 18 of the 27 included cross-sectional studies had a low risk of bias. The remaining nine studies had a moderate risk of bias. An overview of the risk of bias assessment is presented in table 1.

**Table 1.** JBI Checklist for Analytical Cross-Sectional Studies.

| Reference | Inclusion criteria | Study subjects and settings | Exposure measurement | Measurement of the condition | Identification of confounding factors | Dealing with confounding factors | Outcome measurement | Statistical analysis | Score | Risk of Bias |
| --- | --- | --- | --- | --- | --- | --- | --- | --- | --- | --- |
| **Normal hearing** |  |  |  |  |  |  |  |  |  |  |
| Barnea et al., 1990 (52) | N | Y | Y | Y | U | U | Y | Y | 5/8 | Moderate |
| Bilgen et al., 2010 (47) | Y | Y | Y | U | Y | N | Y | Y | 6/8 | Moderate |
| Cartocci et al., 2012 (53) | Y | Y | Y | Y | Y | Y | Y | Y | 8/8 | Low |
| Dadoo et al., 2019 (54) | Y | Y | Y | Y | Y | Y | Y | Y | 8/8 | Low |
| Dos Santos Filha et al., 2014 (55) | Y | Y | Y | Y | Y | Y | Y | Y | 8/8 | Low |
| Dos Santos Filha et al., 2015 (16) | Y | Y | Y | Y | Y | Y | Y | Y | 8/8 | Low |
| Gabr et al., 2020 (56) | Y | Y | Y | Y | Y | Y | Y | Y | 8/8 | Low |
| Guest et al., 2017 (49) | Y | Y | Y | Y | Y | Y | Y | Y | 8/8 | Low |
| Hsu et al., 2013 (57) | Y | Y | Y | U | N | U | Y | Y | 5/8 | Moderate |
| Kehrle et al., 2008 (58) | Y | Y | Y | U | Y | Y | Y | Y | 7/8 | Low |
| Konadath et al., 2016 (59) | Y | Y | Y | Y | N | N | U | Y | 5/8 | Moderate |
| Makar et al., 2017 (60) | Y | Y | U | Y | Y | Y | U | Y | 6/8 | Moderate |
| Nemati et al., 2014 (61) | Y | Y | Y | U | Y | Y | Y | Y | 7/8 | Low |
| Omidvar et al., 2018 (51) | Y | Y | Y | Y | Y | Y | Y | Y | 8/8 | Low |
| Paul et al., 2017 (50) | Y | Y | Y | Y | U | U | Y | Y | 6/8 | Moderate |
| Schaette et al., 2011 (62) | U | Y | Y | Y | Y | Y | Y | Y | 7/8 | Low |
| Shim et al., 2017 (63) | Y | Y | Y | U | Y | Y | Y | Y | 7/8 | Low |
| Shim et al., 2021 (64) | Y | Y | Y | Y | Y | Y | Y | Y | 8/8 | Low |
| Song et al., 2018 (65) | Y | Y | Y | Y | U | U | Y | Y | 6/8 | Moderate |
| Theodoroff et al., 2011 (48) | Y | Y | Y | Y | Y | Y | Y | Y | 8/8 | Low |
| **Mixed population** |  |  |  |  |  |  |  |  |  |  |
| Gilles et al., 2016 (43) | Y | Y | Y | Y | Y | Y | Y | Y | 8/8 | Low |
| Gu et al., 2012 (66) | Y | Y | Y | Y | Y | Y | Y | Y | 8/8 | Low |
| Ikner et al., 1990 (67) | Y | Y | U | N | Y | Y | Y | Y | 6/8 | Moderate |
| **Hearing loss** |  |  |  |  |  |  |  |  |  |  |
| Attias et al., 1993 (68) | U | Y | Y | Y | Y | Y | Y | Y | 7/8 | Low |
| Attias et al., 1996 (69) | U | Y | Y | Y | Y | Y | Y | Y | 7/8 | Low |
| Pinkl et al., 2017 (46) | Y | Y | Y | Y | Y | Y | Y | N | 7/8 | Low |
| Rosenhall et al., 1995 (70) | Y | Y | Y | Y | U | U | Y | U | 5/8 | Moderate |
| Y = yes<br>N = no<br>U = unclear |  |  |  |  |  |  |  |  |  |  |

### Auditory brainstem responses (ABRs)

Results of the 24 cross-sectional studies that investigated ABR latencies and amplitudes are summarized in the appendix (S4 table). Results for tinnitus patients with and without hearing loss will be discussed separately in the sections below.

#### Tinnitus patients with hearing loss: best-evidence synthesis

Due to the clinical heterogeneity between studies investigating ABRs in tinnitus patients with hearing loss, statistical pooling was not feasible. Therefore, a best-evidence synthesis (71) was performed. The standardized mean differences presented in the included studies are shown in figure 2.

**Figure 2.**
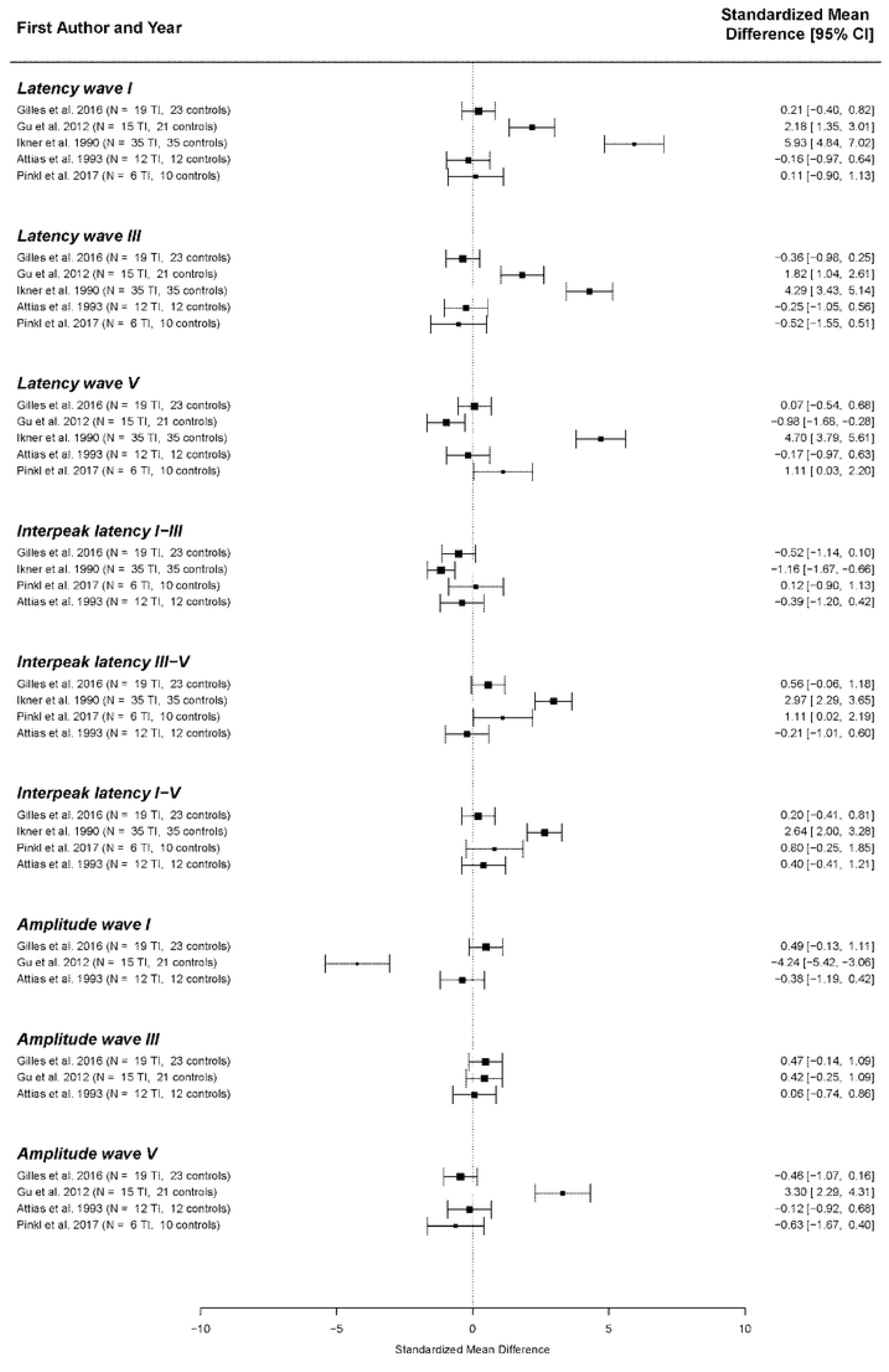
Standardized mean differences for the different ABR components across studies. The studies of Attias et al. 1996 (69) and Rosenhall et al. (70) could not be included in this analysis, since numerical results of different ABR components were not reported in these papers.

The overall results of the best-evidence synthesis show that no consistent changes in any of the ABR components were present in tinnitus patients with hearing loss. There is a possible weak tendency toward longer latencies of waves I, III, and V. However, these results are heavily influenced by an outlier (67). Furthermore, a very subtle tendency toward a shorter IPL I-III and longer IPL III-V and I-V are shown. Regarding ABR amplitudes, no consistent differences could be identified.

#### Tinnitus patients without hearing loss: meta-analysis

Eleven studies investigating ABR components in normal hearing tinnitus patients were included in the meta-analysis. A detailed overview of the reasons for exclusion in the final meta-analysis can be found in the appendix (S6 table). The characteristics of the study participants of the studies included in our meta-analysis are shown in the appendix (S7 table).

The following ABR components were included in data pooling: latencies of waves I (n = 9), III (n = 9), and V (n = 10); interpeak latencies (IPLs) I-III (n = 7), III-V (n = 8), and I-V (n = 7), amplitude wave I (n = 3) and V (n = 3). Standardized Mean Differences (SMDs) between tinnitus patients and controls within each study were calculated for these elements. The final multivariate model, shown in figure 3, resulted in significant SMDs between tinnitus patients and controls for four of the included ABR components. Latencies of waves I (SMD = 0.66 ms, p < 0.001), III (SMD = 0.43 ms p < 0.001), and V (SMD = 0.47 ms, p < 0.01) are shown to be significantly longer in tinnitus patients than controls. Statistical heterogeneity for wave I amplitude was too high (I^2^ = 89.84%), so data could not be pooled. SMDs for interpeak latencies I-III, III-V, and I-V and amplitude of wave V were close to zero.

**Figure 3.**
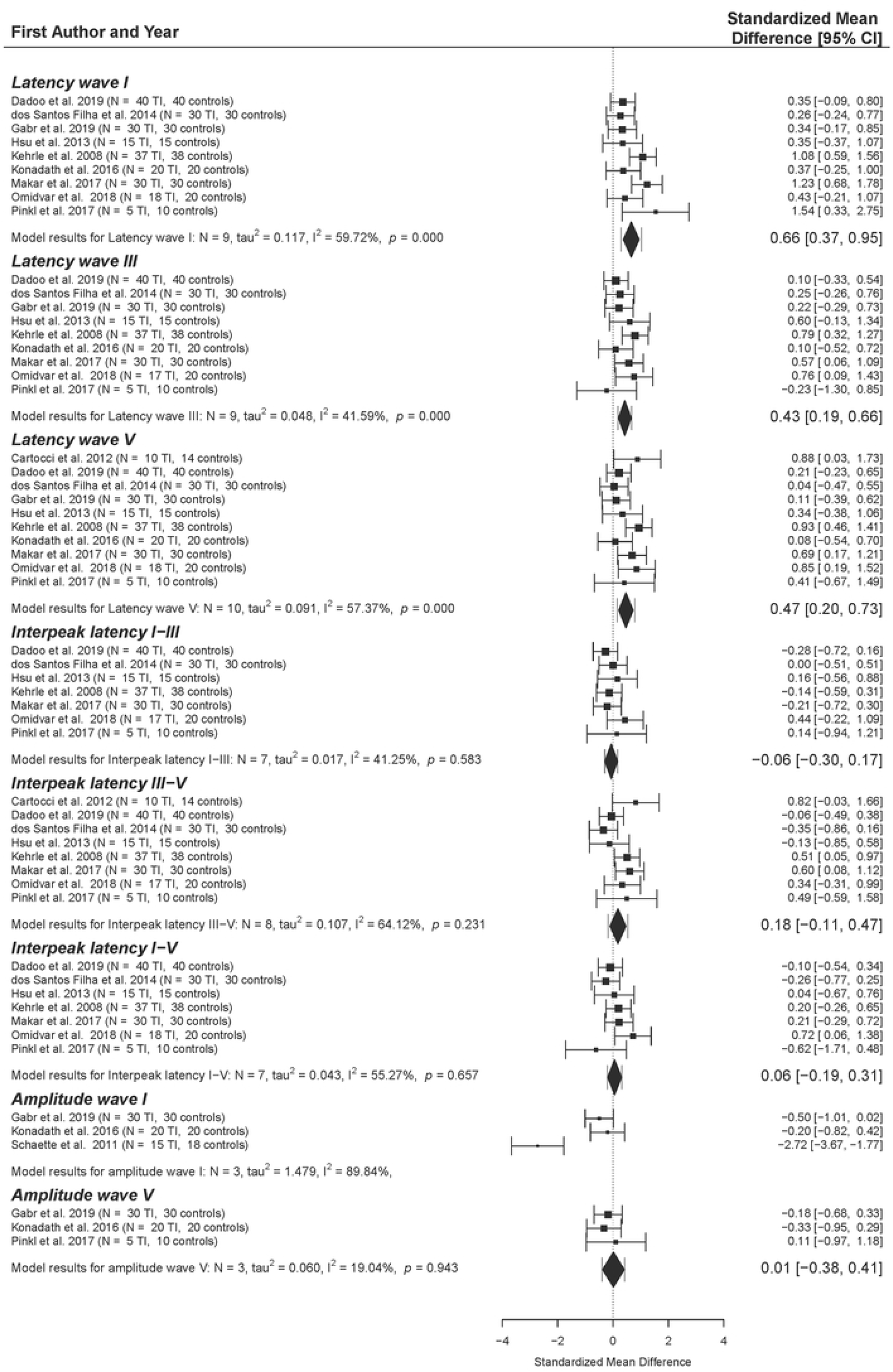
Forest plot of the primary multivariate analysis. Results are grouped according to ABR component. Results from individual papers are presented as Standardized Mean Differences (SMD) ± 95% confidence intervals. Overall results from the primary meta-analytic model are given for each component. SMD with 95% confidence intervals are represented by diamonds, while error bars correspond to credibility/prediction intervals, defined as the intervals where approximately 95% of the true outcomes are expected to fall.

For each component, post hoc analyses were performed by excluding possible outliers or influencing studies. This is discussed in detail in section 8 in the appendix (S8). Overall, the removal of outliers and influential papers did not change the outcomes compared to the primary analyses for all ABR components included in the meta-analyses.

Publication bias was investigated using funnel plots and Egger’s regression tests for each ABR component separately. No evidence for publication bias was found for any of the other ABR components. Funnel plots for ABR latencies of waves I, III, and V, as well as forest plots with the outliers and influential papers excluded, are given in the appendix (S8 and S9 figures).

### Middle-latency responses (MLRs)

MLR latencies and amplitudes were investigated in three studies, whose results are depicted in the appendix (S5 table). Regarding Na and Pa latencies, none of these studies reported significant differences between tinnitus patients with normal hearing and controls. Not all possible MLR waves were examined in all four of these papers. For instance, wave Pb latency was discussed in only two of them. No consistent differences in any of the other MLR latencies or amplitudes could be identified.

### Frequency-following responses (FFRs)

Being only investigated by three of the included studies, the FFR was the least studied AEP in our systematic review. More specifically, Guest et al. (49), Paul et al. (50), and Omidvar et al. (51) examined the fundamental frequency (F_0_) in tinnitus patients with normal hearing compared to controls. All three of these studies reported lower, though non-significant, response amplitudes in tinnitus patients. However, it must be noted that all of these studies used different stimuli and intensity levels to elicit the FFR (49-51). By eliciting the FFR with a 40 ms synthesized syllable /da/, Omidvar et al. (51) also reported significantly decreased amplitudes of the first formant frequency range (F_1_) and higher frequency region (HH) in tinnitus patients. Moreover, the mean latencies of all FFR waves (more specifically, waves V, A, C, D, E, F, and O) were significantly longer in subjects with tinnitus than in the control group.

## Discussion

Our meta-analysis showed prolonged latencies of waves I, III, and V in tinnitus patients with normal hearing. The best-evidence synthesis in tinnitus patients with hearing loss did not reveal any consistent differences.

In contrast to our expectations of reduced wave latencies due to increased spontaneous firing rates and neural synchrony, our meta-analyses revealed consistent prolongation of wave latencies in several studies.

A prolongation of the latency of wave I, parallel to a lengthening of the later ABR latencies of waves III and V, occurs in ears with sensorineural hearing loss (72-75). No differences in interpeak latencies were found, which further supports this theory (74, 75). Thus, it suggests that patients of the tinnitus group might have had sensorineural hearing loss at higher frequencies which cannot be measured by click ABR (58, 60). In addition, in normal hearing tinnitus patients somatosensory triggers such as temporomandibular dysfunction could also modulate auditory brainstem activity causing delayed ABR latencies (13, 76, 77).

The mean age over all studies in our meta-analysis was almost 5 years higher for tinnitus patients (38.9 years) compared to controls (34.1 years). This difference could be the cause of a small age bias, which might also influence the results. The possibility of a gender bias is rather small, since there only was a minor difference in mean proportion of genders between tinnitus (proportion of males = 0.61) and controls (proportion of males = 0.59).

In previous research, a decreased amplitude of wave I has been observed (78, 79). This decrease in amplitude was hypothesized to be caused by the presence of hidden hearing loss, or cochlear synaptopathy, which describes the degeneration of the cochlear synapses without loss of hair cells (51, 62, 80). Our meta-analysis did not replicate these results. However, some researchers argue that the click ABR is not sensitive enough to identify cochlear synaptopathy in humans (49). Thus, this theory of cochlear synaptopathy in tinnitus patients also cannot be refuted by our results. In the study by Guest et al. (49), FFRs were also acquired in order to examine the presence of cochlear synaptopathy in tinnitus patients. More specifically, fundamental frequency (F_0_) differences were expected to increase due to synaptopathy. However, no significant effects were found.

### Clinical implications

In the review by Cardon et al. (33), the parietocentral (34) P300 is put forward as a potential biomarker for tinnitus at cortical level. The current review proves that by acquiring ABR waves I, III, and V, changes earlier on in the auditory pathway, more specifically at brainstem level, can be revealed in some tinnitus patients. At present, we cannot confirm whether the cortical changes are a result of the changes earlier on in the auditory pathway. Moreover, the P300 depends on the processing of the stimulus context and levels of attention and arousal (36), and is therefore often used as a measure of cognitive processing (35). In contrast, ABR waves are unaffected by arousal and attention (81, 82), therefore providing us with different information on auditory processing. Thus, auditory brainstem responses and cortical auditory evoked potentials might complement each other to identify the various changes on different levels of the auditory pathway in tinnitus patients with identical or different underlying pathologies.

Furthermore, even though tinnitus patients can present with normal hearing, reflected by a normal pure tone audiometry, sensorineural hearing loss at high frequencies could still be present. For that reason, it may be of interest to acquire ABRs in tinnitus patients who present with a normal audiogram anyway and to perform a high frequency audiometry, in order to diagnose potential latency shifts associated with high-frequency hearing loss.

### Directions for further research

Risk of bias assessment revealed a low risk of bias in the majority of the included studies. Throughout the various studies, identifying and dealing with confounding factors proved to be the most common source of risk of bias. However, it is well known that AEPs can be affected by several factors, including age, gender, and hearing loss (83-85). For instance, several of the papers in the current review did not report age or gender of participants (46, 52, 61, 67), or did not mention whether matching was performed (16, 47, 48, 50, 54, 59, 66). Therefore, we strongly recommend future research to identify and report these confounding factors, and to set clear inclusion criteria accordingly to avoid sampling errors.

We were not able to draw any conclusions on possible differences in MLR and FFR potentials, mainly because insufficient studies investigating these components could be included in our systematic review. Since our review was able to highlight AEP changes at brainstem level and the review by Cardon et al. (33) did so for the cortical level, there still remains a knowledge gap about whether changes occur at subcortical level. Since MLRs are considered to represent subcortical activation (22) and FFRs arise from multiple cortical and subcortical sources (25, 86), these potentials might help to fill in this knowledge gap. This would allow us to further understand which changes occur in tinnitus patients along the complete auditory pathway, from cochlea to cortex. Thus, our recommendation is to conduct cross-sectional studies measuring MLRs, and FFRs, which are carried out in sufficiently large and homogeneous samples.

### Strengths and limitations

To our knowledge, this is the first systematic review and meta-analysis investigating both short- and middle-latency AEPs in tinnitus patients. The use of a powerful and well-constructed methodology contributed to the strength of the present paper. More specifically, risk of bias assessment was performed by two independent reviewers, a broad search strategy was constructed, and this paper was reported according to the PRISMA guidelines (39).

Nevertheless, we encountered a few limitations. Although we intended to homogenize the included data in our meta-analysis as much as possible, some clinical heterogeneity is inevitable. For instance, there were some differences in gender ratio and mean age across studies. Some variation in the methodology for the acquisition ABRs was also present, such as the ABR system, the type of transducer, the presentation level, and the filtering settings. Moreover, some papers that were eligible to be included in our meta-analyses did not report ABR latencies and amplitudes, and consequently could not be included in the final analyses.

Additionally, most papers did not provide many details on the tinnitus characteristics of the subjects. These include duration, loudness, and subjective severity of tinnitus.

## Conclusion

Significantly longer latencies of ABR waves I, III, and V are shown in tinnitus patients with normal hearing compared to controls. This could be explained by a high frequency sensorineural hearing loss or other less known modulating factors such as cochlear synaptopathy or somatosensory tinnitus generators. No conclusions on possible changes at subcortical level could be drawn yet.

## Data Availability

All relevant data are within the manuscript and its Supporting Information files.

## Supporting information captions

**S1 Appendix**. Appendix to the main manuscript, containing the full data extraction tables.

**S2 File**. Prisma checklist.

---

**Figure S1.**
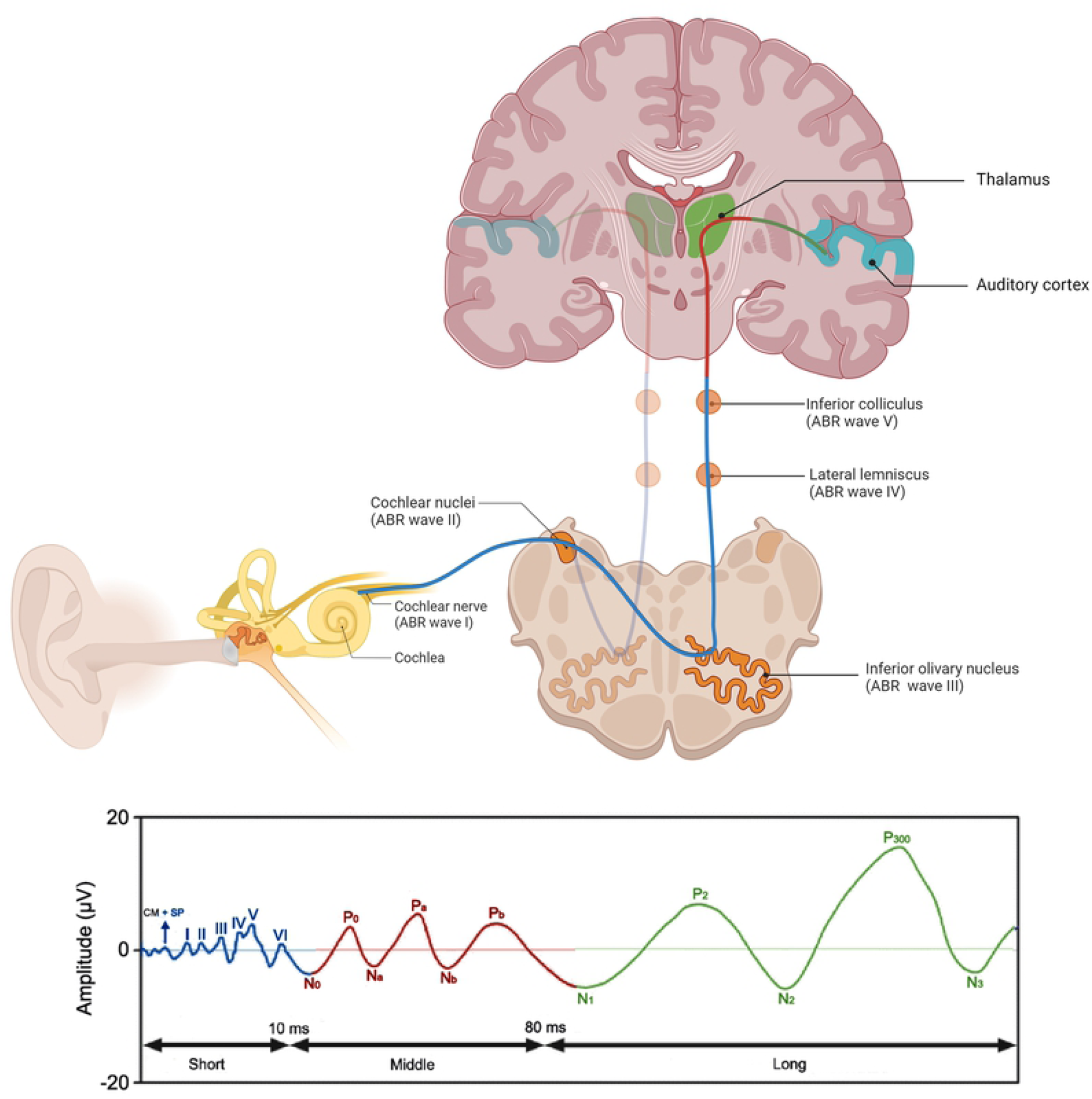

**Figure S8.**
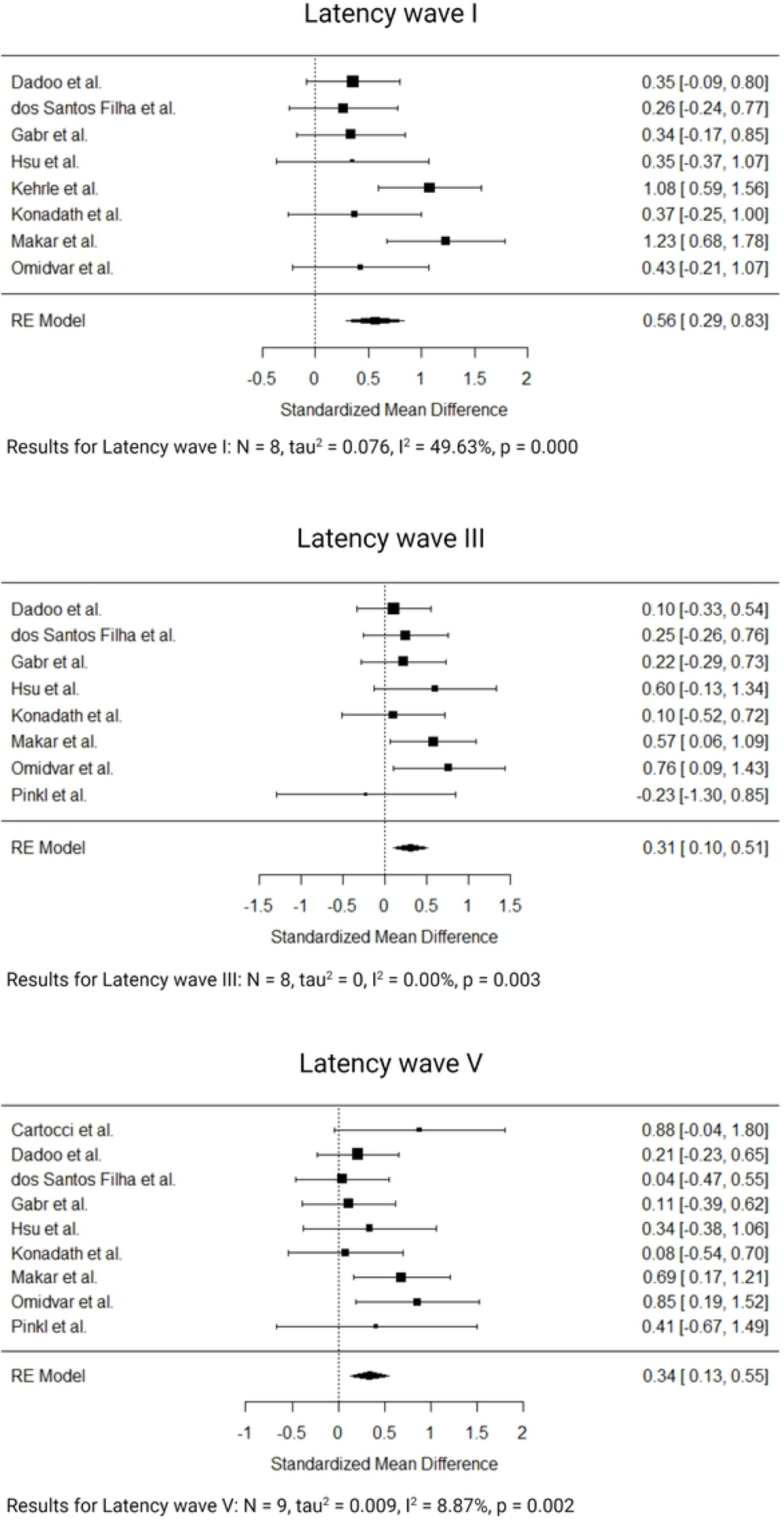

**Figure S9.**
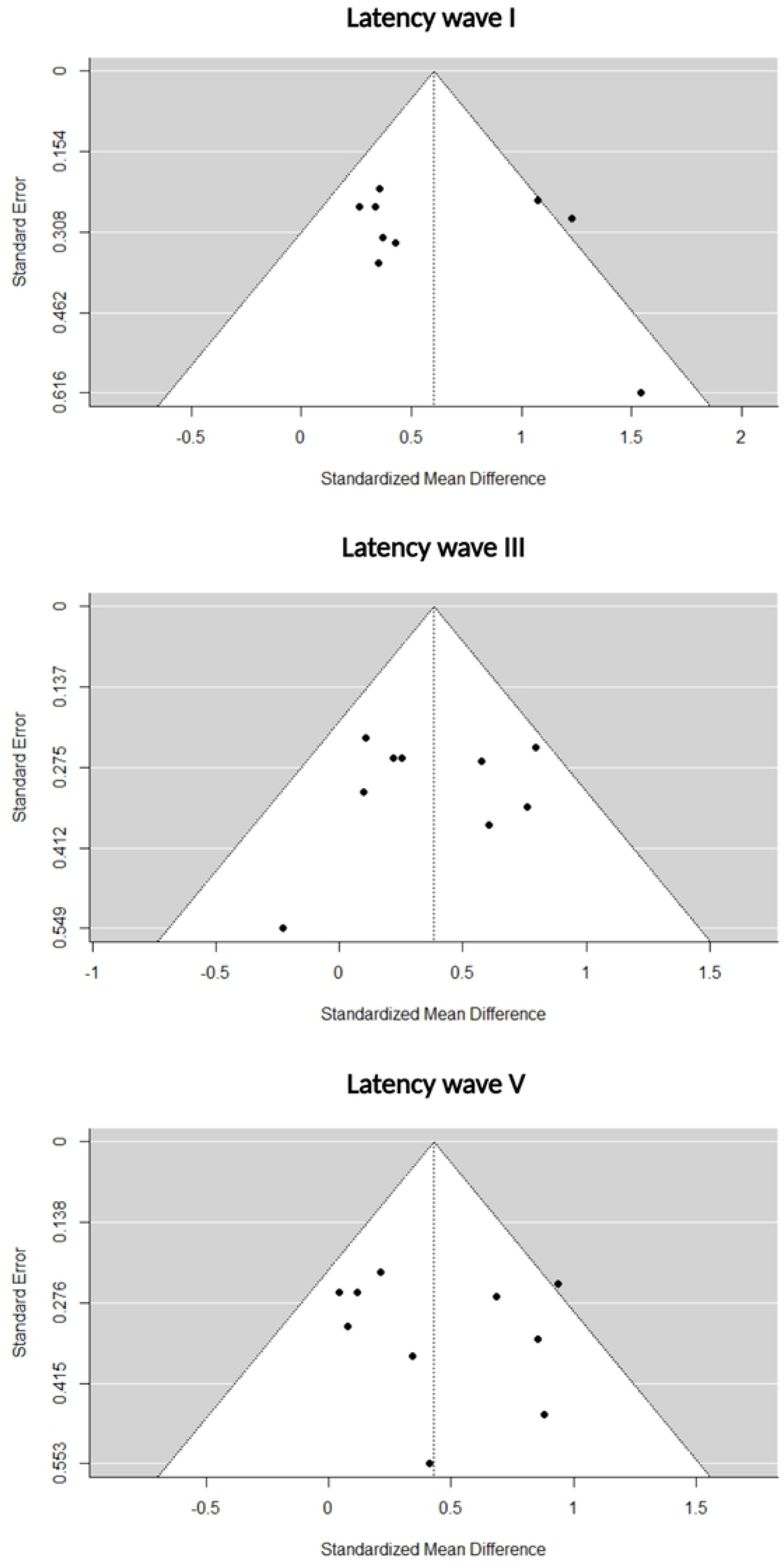

## References

1. Baguley D, McFerran D, Hall D. Tinnitus. The Lancet. 2013;382(9904):1600–7.

2. Tyler RS, Baker LJ. Difficulties experienced by tinnitus sufferers. J Speech Hear Disord. 1983;48(2):150–4.

3. Reed GF. An audiometric study of two hundred cases of subjective tinnitus. AMA Arch Otolaryngol. 1960;71:84–94.

4. Vernon J. Attemps to relieve tinnitus. J Am Audiol Soc. 1977;2(4):124–31.

5. Theodoroff SM, Kaltenbach JA. The Role of the Brainstem in Generating and Modulating Tinnitus. Am J Audiol. 2019;28(1s):225–38.

6. Møller AR. Sensorineural Tinnitus: Its Pathology and Probable Therapies. Int J Otolaryngol. 2016;2016:2830157.

7. Haider HF, Bojić T, Ribeiro SF, Paço J, Hall DA, Szczepek AJ. Pathophysiology of Subjective Tinnitus: Triggers and Maintenance. Front Neurosci. 2018;12:866.

8. Kaltenbach JA, Godfrey DA, Neumann JB, McCaslin DL, Afman CE, Zhang J. Changes in spontaneous neural activity in the dorsal cochlear nucleus following exposure to intense sound: relation to threshold shift. Hear Res. 1998;124(1-2):78–84.

9. Kaltenbach JA, Afman CE. Hyperactivity in the dorsal cochlear nucleus after intense sound exposure and its resemblance to tone-evoked activity: a physiological model for tinnitus. Hear Res. 2000;140(1-2):165–72.

10. Brozoski TJ, Bauer CA, Caspary DM. Elevated fusiform cell activity in the dorsal cochlear nucleus of chinchillas with psychophysical evidence of tinnitus. J Neurosci. 2002;22(6):2383–90.

11. Kaltenbach JA, Zacharek MA, Zhang J, Frederick S. Activity in the dorsal cochlear nucleus of hamsters previously tested for tinnitus following intense tone exposure. Neurosci Lett. 2004;355(1-2):121–5.

12. van Gendt MJ, Boyen K, de Kleine E, Langers DR, van Dijk P. The relation between perception and brain activity in gaze-evoked tinnitus. J Neurosci. 2012;32(49):17528–39.

13. Lanting CP, de Kleine E, Eppinga RN, van Dijk P. Neural correlates of human somatosensory integration in tinnitus. Hear Res. 2010;267(1-2):78–88.

14. Lanting CP, de Kleine E, van Dijk P. Neural activity underlying tinnitus generation: results from PET and fMRI. Hear Res. 2009;255(1-2):1–13.

15. Melcher JR, Sigalovsky IS, Guinan JJ, Levine RA. Lateralized tinnitus studied with functional magnetic resonance imaging: Abnormal inferior colliculus activation. Journal of Neurophysiology. 2000;83(2):1058–72.

16. dos Santos Filha VA, Samelli AG, Matas CG. Middle Latency Auditory Evoked Potential (MLAEP) in Workers with and without Tinnitus who are Exposed to Occupational Noise. Med Sci Monit. 2015;21:2701–6.

17. Radeloff A, Cebulla M, Shehata-Dieler W. [Auditory evoked potentials: basics and clinical applications]. Laryngorhinootologie. 2014;93(9):625–37.

18. Alain C, Roye A, Arnott SR. Middle-and long-latency auditory evoked potentials: What are they telling us on central auditory disorders? Handbook of clinical neurophysiology. 2013:177–99.

19. De Cosmo G, Aceto P, Clemente A, Congedo E. Auditory evoked potentials. Minerva Anestesiol. 2004;70(5):293–7.

20. Burkard R, Eggermont J, Don M. Auditory evoked potentials: basic principles and clinical application: Lippincott Williams & Wilkins.; 2007.

21. Lammers M. Auditory pathway functioning in prelingual deafness. The clinical consequences for cochlear implantation. Utrecht, the Netherlands: University Medical Center Utrecht; 2015.

22. Crivelli D, Balconi M. Event-Related Electromagnetic Responses☆. Reference Module in Neuroscience and Biobehavioral Psychology: Elsevier; 2017.

23. Krizman J, Kraus N. Analyzing the FFR: A tutorial for decoding the richness of auditory function. Hear Res. 2019;382:107779.

24. Kraus N, Anderson S, White-Schwoch T. The Frequency-Following Response: A Window into Human Communication. In: Kraus N, Anderson S, White-Schwoch T, Fay RR, Popper AN, editors. The Frequency-Following Response: A Window into Human Communication. Cham: Springer International Publishing; 2017. p. 1–15.

25. Coffey EBJ, Nicol T, White-Schwoch T, Chandrasekaran B, Krizman J, Skoe E, et al. Evolving perspectives on the sources of the frequency-following response. Nat Commun. 2019;10(1):5036.

26. Bidelman GM. Multichannel recordings of the human brainstem frequency-following response: Scalp topography, source generators, and distinctions from the transient ABR. Hearing Research. 2015;323:68–80.

27. Chandrasekaran B, Kraus N. The scalp-recorded brainstem response to speech: neural origins and plasticity. Psychophysiology. 2010;47(2):236–46.

28. Sohmer H, Pratt H, Kinarti R. Sources of frequency following responses (FFR) in man. Electroencephalogr Clin Neurophysiol. 1977;42(5):656–64.

29. White-Schwoch T, Nicol T, Warrier CM, Abrams DA, Kraus N. Individual differences in human auditory processing: insights from single-trial auditory midbrain activity in an animal model. Cerebral Cortex. 2017;27(11):5095–115.

30. White-Schwoch T, Anderson S, Krizman J, Nicol T, Kraus N. Case studies in neuroscience: subcortical origins of the frequency-following response. J Neurophysiol. 2019;122(2):844–8.

31. Malmierca MS. Anatomy and Physiology of the Mammalian Auditory System. In: Jaeger D, Jung R, editors. Encyclopedia of Computational Neuroscience. New York, NY: Springer New York; 2015. p. 155–86.

32. Schochat E, Rocha-Muniz CN, Filippini R. Understanding Auditory Processing Disorder Through the FFR. The Frequency-Following Response 2017. p. 225–50.

33. Cardon E, Joossen I, Vermeersch H, Jacquemin L, Mertens G, Vanderveken OM, et al. Systematic review and meta-analysis of late auditory evoked potentials as a candidate biomarker in the assessment of tinnitus. Plos One. 2020;15(12).

34. Linden DE. The p300: where in the brain is it produced and what does it tell us? Neuroscientist. 2005;11(6):563–76.

35. Reinvang I. Cognitive event-related potentials in neuropsychological assessment. Neuropsychol Rev. 1999;9(4):231–48.

36. Polich J, Howard L, Starr A. P300 latency correlates with digit span. Psychophysiology. 1983;20(6):665–9.

37. Domarecka E, Olze H, Szczepek AJ. Auditory Brainstem Responses (ABR) of Rats during Experimentally Induced Tinnitus: Literature Review. Brain Sciences. 2020;10(12).

38. Moher D, Shamseer L, Clarke M, Ghersi D, Liberati A, Petticrew M, et al. Preferred reporting items for systematic review and meta-analysis protocols (PRISMA-P) 2015 statement. Syst Rev. 2015;4(1):1.

39. Page MJ, McKenzie JE, Bossuyt PM, Boutron I, Hoffmann TC, Mulrow CD, et al. The PRISMA 2020 statement: An updated guideline for reporting systematic reviews. PLoS Med. 2021;18(3):e1003583.

40. Morton S, Berg A, Levit L, Eden J. Finding what works in health care: standards for systematic reviews. 2011.

41. Marshall CA, Boland L, Westover LA, Wickett S, Roy L, Mace J, et al. Occupational experiences of homelessness: A systematic review and meta-aggregation. Scand J Occup Ther. 2020;27(6):394–407.

42. Viechtbauer W. Conducting meta-analyses in R with the metafor package. Journal of statistical software. 2010;36(3):1–48.

43. Gilles A, Schlee W, Rabau S, Wouters K, Fransen E, Van de Heyning P. Decreased Speech-In-Noise Understanding in Young Adults with Tinnitus. Front Neurosci. 2016;10:288.

44. Jackson D, White IR, Riley RD. Quantifying the impact of between-study heterogeneity in multivariate meta-analyses. Stat Med. 2012;31(29):3805–20.

45. Light RJ, Pillemer DB. Summing up: the science of reviewing research. 1984.

46. Pinkl J, Wilson MJ, Billingsly D, Munguia-Vazquez R. Detailed Analysis of High Frequency Auditory Brainstem Response in Patients with Tinnitus: A Preliminary Study. Int Tinnitus J. 2017;21(1):35–43.

47. Bilgen C, Sezer B, Kirazli T, Gunbay T. Tinnitus in Temporomandibular Disorders: Electrophysiological Aspects. Journal of International Advanced Otology. 2010;6(2):167–72.

48. Theodoroff S, Chambers R, McMillan R. Auditory middle latency responses in individuals with debilitating tinnitus. Int Tinnitus J. 2011;16(2):104–10.

49. Guest H, Munro KJ, Prendergast G, Howe S, Plack CJ. Tinnitus with a normal audiogram: Relation to noise exposure but no evidence for cochlear synaptopathy. Hear Res. 2017;344:265–74.

50. Paul BT, Bruce IC, Roberts LE. Evidence that hidden hearing loss underlies amplitude modulation encoding deficits in individuals with and without tinnitus. Hear Res. 2017;344:170–82.

51. Omidvar S, Mahmoudian S, Khabazkhoob M, Ahadi M, Jafari Z. Tinnitus Impacts on Speech and Non-speech Stimuli. Otol Neurotol. 2018;39(10):e921–e8.

52. Barnea G, Attias J, Gold S, Shahar A. Tinnitus with normal hearing sensitivity: extended high-frequency audiometry and auditory-nerve brain-stem-evoked responses. Audiology. 1990;29(1):36–45.

53. Cartocci G, Attanasio G, Fattapposta F, Locuratolo N, Mannarelli D, Filipo R. An electrophysiological approach to tinnitus interpretation. Int Tinnitus J. 2012;17(2):152–7.

54. Dadoo S, Sharma R, Sharma V. Oto-acoustic emissions and brainstem evoked response audiometry in patients of tinnitus with normal hearing. Int Tinnitus J. 2019;23(1):17–25.

55. dos Santos-Filha VAV, Samelli AG, Matas CG. Noise-induced tinnitus: auditory evoked potential in symptomatic and asymptomatic patients. Clinics. 2014;69(7):487–90.

56. Gabr TA, Lasheen RM. Binaural Interaction in Tinnitus Patients. Audiol Neurootol. 2020;25(6):315–22.

57. Hsu SY, Wang PC, Yang TH, Lin TF, Hsu SH, Hsu CJ. Auditory efferent dysfunction in normal-hearing chronic idiopathic tinnitus. B-ent. 2013;9(2):101–9.

58. Kehrle HM, Granjeiro RC, Sampaio AL, Bezerra R, Almeida VF, Oliveira CA. Comparison of auditory brainstem response results in normal-hearing patients with and without tinnitus. Arch Otolaryngol Head Neck Surg. 2008;134(6):647–51.

59. Konadath S, Manjula P. Auditory brainstem response and late latency response in individuals with tinnitus having normal hearing. Intractable Rare Dis Res. 2016;5(4):262–8.

60. Makar SK, Mukundan G, Gore G. Auditory System Synchronization and Cochlear Function in Patients with Normal Hearing With Tinnitus: Comparison of Multiple Feature with Longer Duration and Single Feature with Shorter Duration Tinnitus. Int Tinnitus J. 2017;21(2):133–8.

61. Nemati S, Faghih Habibi A, Panahi R, Pastadast M. Cochlear and brainstem audiologic findings in normal hearing tinnitus subjects in comparison with non-tinnitus control group. Acta Med Iran. 2014;52(11):822–6.

62. Schaette R, McAlpine D. Tinnitus with a normal audiogram: physiological evidence for hidden hearing loss and computational model. J Neurosci. 2011;31(38):13452–7.

63. Shim HJ, An YH, Kim DH, Yoon JE, Yoon JH. Comparisons of auditory brainstem response and sound level tolerance in tinnitus ears and non-tinnitus ears in unilateral tinnitus patients with normal audiograms. PLoS One. 2017;12(12):e0189157.

64. Shim HJ, Cho YT, Oh HS, An YH, Kim DH, Kang YS. Within-Subject Comparisons of the Auditory Brainstem Response and Uncomfortable Loudness Levels in Ears With and Without Tinnitus in Unilateral Tinnitus Subjects With Normal Audiograms. Otol Neurotol. 2021;42(1):10–7.

65. Song K, Shin SA, Chang DS, Lee HY. Audiometric Profiles in Patients With Normal Hearing and Bilateral or Unilateral Tinnitus. Otol Neurotol. 2018;39(6):e416–e21.

66. Gu JW, Herrmann BS, Levine RA, Melcher JR. Brainstem auditory evoked potentials suggest a role for the ventral cochlear nucleus in tinnitus. J Assoc Res Otolaryngol. 2012;13(6):819–33.

67. Ikner CL, Hassen AH. The effect of tinnitus on ABR latencies. Ear Hear. 1990;11(1):16–20.

68. Attias J, Urbach D, Gold S, Shemesh Z. Auditory event related potentials in chronic tinnitus patients with noise induced hearing loss. Hear Res. 1993;71(1-2):106–13.

69. Attias J, Pratt H, Reshef I, Bresloff I, Horowitz G, Polyakov A, et al. Detailed analysis of auditory brainstem responses in patients with noise-induced tinnitus. Audiology. 1996;35(5):259–70.

70. Rosenhall U, Axelsson A. Auditory brainstem response latencies in patients with tinnitus. Scand Audiol. 1995;24(2):97–100.

71. Slavin RE. Best evidence synthesis: an intelligent alternative to meta-analysis. J Clin Epidemiol. 1995;48(1):9–18.

72. Coats AC, Martin JL. Human Auditory Nerve Action Potentials and Brain Stem Evoked Responses: Effects of Audiogram Shape and Lesion Location. Archives of Otolaryngology. 1977;103(10):605–22.

73. Watson DR. The effects of cochlear hearing loss, age and sex on the auditory brainstem response. Audiology. 1996;35(5):246–58.

74. Yilmaz MS, Guven M, Cesur S, Oguz H. The auditory brainstem responses in patients with unilateral cochlear hearing loss. Indian journal of otolaryngology and head and neck surgery : official publication of the Association of Otolaryngologists of India. 2013;65(3):203–9.

75. Lin HC, Chou YC, Wang CH, Hung LW, Shih CP, Kang BH, et al. Correlation between auditory brainstem response and hearing prognosis in idiopathic sudden sensorineural hearing loss patients. Auris Nasus Larynx. 2017;44(6):678–84.

76. Balmer TS, Trussell LO. Trigeminal Contributions to the Dorsal Cochlear Nucleus in Mouse. Front Neurosci. 2021;15:715954.

77. Shore SE. Multisensory integration in the dorsal cochlear nucleus: unit responses to acoustic and trigeminal ganglion stimulation. Eur J Neurosci. 2005;21(12):3334–48.

78. Milloy V, Fournier P, Benoit D, Norena A, Koravand A. Auditory Brainstem Responses in Tinnitus: A Review of Who, How, and What? Frontiers in Aging Neuroscience. 2017;9.

79. Chen F, Zhao F, Mahafza N, Lu W. Detecting Noise-Induced Cochlear Synaptopathy by Auditory Brainstem Response in Tinnitus Patients With Normal Hearing Thresholds: A Meta-Analysis. Front Neurosci. 2021;15:778197.

80. Liberman MC, Kujawa SG. Cochlear synaptopathy in acquired sensorineural hearing loss: Manifestations and mechanisms. Hear Res. 2017;349:138–47.

81. Picton TW, Hillyard SA, Galambos R, Schiff M. Human auditory attention: a central or peripheral process? Science. 1971;173(3994):351–3.

82. Picton TW, Hillyard SA. Human auditory evoked potentials. II. Effects of attention. Electroencephalogr Clin Neurophysiol. 1974;36(2):191–9.

83. Konrad-Martin D, Dille MF, McMillan G, Griest S, McDermott D, Fausti SA, et al. Age-related changes in the auditory brainstem response. J Am Acad Audiol. 2012;23(1):18–35; quiz 74-5.

84. Mitchell C, Phillips DS, Trune DR. Variables affecting the auditory brainstem response: audiogram, age, gender and head size. Hear Res. 1989;40(1-2):75–85.

85. Jerger J, Johnson K. Interactions of age, gender, and sensorineural hearing loss on ABR latency. Ear Hear. 1988;9(4):168–76.

86. Aiken SJ, Picton TW. Envelope and spectral frequency-following responses to vowel sounds. Hear Res. 2008;245(1-2):35–47.

